# Re-opening schools in a context of low COVID-19 contagion: Consequences for teachers, students and their parents*

**DOI:** 10.1101/2021.03.25.21254219

**Authors:** Anna Godøy, Maja Weemes Grøtting, Rannveig Kaldager Hart

## Abstract

To balance children’s right to schooling with contagion management, knowing how school re-openings affect the spread of COVID-19 is crucial. This paper considers effects on testing and positive tests for COVID-19 of re-opening Norwegian schools after a six-week closure to reduce contagion. We estimate the effect of school reopening for teachers, parents and students using an event study/difference-in-differences design with comparison groups with minimal exposure to in-person schooling. We find no evidence that incidence increased following re-opening for either students, parents, or teachers pooled across grade levels. We find some suggestive evidence that infection rates among upper secondary school teachers increased; however, the effects are small and transitory. At low levels of contagion, schools can safely be re-opened when other social distancing policies remain in place.

## 1 Introduction

During the spring of 2020 schools across the world were closed as part of an effort to curb the first wave of the COVID-19 pandemic. A school closure of this dimension is unprecedented in modern history (Insights For Education 2020). When instruction is given online, lack of in-person schooling can have negative impacts on children’s development, health and well-being (see, e.g., Donohue & Miller 2020, Dooley et al. 2020). Early evidence suggests a “COVID learning gap”, with increased social disparities in learning outcomes (Engzell et al. 2021), possibly driven by parents with higher social background giving more extensive support to online learning (Bacher-Hicks et al. 2020). Moreover, the value of lost lifetime income due to reduced human capital and reduced parental labor supply is estimated to NOK 1.7 billion (200 million USD) each day schools remain closed in Norway (Andresen et al. 2020). Studies from Japan suggest that school closures can have deterimental effects on the health of mothers (Yamamura & Tsustsui 2021) and children (Takaku & Yokoyama 2020). Due to their potentially detrimental consequences, school closures have been among the most controversial virus containment policies. To weigh children’s burdens from school closures against increased contagion risk, policymakers need precise knowledge of how school closures and re-openings affect contagion rates for teachers, students and parents.

In this paper, we assess the effects of school re-openings on the number of tests and confirmed incidence of COVID-19 for teachers, students and parents. While the initial school closures were implemented as part of a broader set of non-pharmaceutical interventions, including the closure of many in-person businesses, the timing of school re-opening did not coincide with any other major policy shifts. School re-openings thus provide an attractive context for isolating the impact of school closure policies.

To identify the effects of re-openings, we implement a difference-in-differences research design, comparing these outcomes before and after schools re-opened across groups with different levels of exposure to in-person schooling. Specifically, we compare outcomes for teachers (grades 1-13) to comparable professionals, high school students (aged 17-19) to young adults aged 20-22, and, finally, parents of high school students (aged 17-19) to parents of young adults aged 20-22. We rely on rich register data covering the universe of Norwegian residents. These data include individual-level data on COVID-19 testing and test results, occupation, and industry as well as other demographic information. We can connect children to parents, enabling comparisons of infection rates and testing among parents with (high) school children to parents of young adult children who recently graduated high school. Similarly, the granular data allow us to identify teachers in elementary and secondary schools as well as comparison groups.

Our main findings can be summarised along the following lines. First, we show that confirmed incidence rates for students, parents and teachers track closely with rates in their respective comparison groups throughout the first wave of the pandemic, including the school reopening. Consistent with this, event study models fail to find evidence that the timing of school re-openings corresponds with a significant shift in confirmed incidence for either students, parents, or teachers. This result holds both in the country as a whole as well as in separate analyses of the capital region with its relatively higher infection incidence. The precision of our estimates allows us to rule out large increases in confirmed incidence following re-opening; e.g. the estimated 95 percent confidence interval for teachers allows us to rule out increases in confirmed incidence larger than 2.1 weekly cases per 100,000 teachers. Overall, our findings suggests that in a context with low contagion rates, where social distancing policies are maintained both in schools and in society as a whole, school re-openings do not necessarily lead to increased incidence among affected groups.

Our findings contribute to the sparse body of knowledge on the impact of school closures on COVID-19 infections. In a recent systematic review, Walsh et al. (2021) identified only ten empirical studies of the effect of school closures on the spread of COVID-19. Effects ranging from no impact to substantial reductions in incidence and mortality were reported. Importantly, for most studies, the effect of school closures could not be isolated. In addition, the studies with the least issues from confounding were among the studies that reported no effects. To our knowledge, only a few examples of quasi-experimental studies aiming at identifying the causal effect of school closures on COVID-19 infection exists. Vlachos et al. (2021) exploit the fact that upper secondary schools in Sweden moved to online teaching, while lower secondary schools did not. They compare teachers and parents of students in the last year of lower secondary school to teachers and parents in the first year of upper secondary school and find somewhat higher infection rates among teachers and parents of the former group.

The studies closest to our study, von Bismarck-Osten et al. (2020) and Isphording et al. (2020), exploit similar variations in school closures and re-openings resulting from staggered summer holidays across German regions. These two studies estimate effects of school summer holidays, finding little evidence that school re-openings increased the spread of COVID-19. Meanwhile, the summer holidays are a period of vast travel in Germany, where people typically travel to regions with higher infection rates (von Bismarck-Osten et al. 2020). Moreover, Germany introduced a policy of testing travellers at an increasing rate during the summer holiday, potentially changing recorded infection rates even when there are no changes in the true underlying incidence. In contrast, our study estimates effects of school re-openings in a context of extremely limited travel. We leverage individual level data on testing and show that there were little differences in testing across our treatment and comparison groups. Finally, while von Bismarck-Osten et al. (2020) and Isphording et al. (2020) analyze data on the county-age group level, we leverage linked individual data on occupation and family linkages to identify teachers and parents, groups that have the highest exposure to in-person schooling. Our findings are also related to the rapidly expanding literature on the effects of other non-pharmaceutical interventions to curb the spread of the virus (Bonacini et al. 2021, Chen et al. 2020, Lyu & Wehby 2020, Qiu et al. 2020, Sears et al. 2020).

More generally, our findings shed light on patterns of COVID-19 transmission among teachers and students. Current research suggests that Norwegian teachers had relatively low contagion levels compared to other professions during the spring of 2020, and somewhat elevated levels during the fall of 2020 (Magnusson et al. 2021). Our approach adds to the understanding of contagion in schools by analysing whether changes in contagion and testing around the time of reopening are larger for teachers than for comparable groups.

Current evidence suggests that severe consequences of COVID-19 disease are rare in children. However, children’s role in the transmission of the virus is still subject to some debate and while symptomatic children are found to shed virus in similar quantities and to infect others in a similar way as adults, it is unclear how infectious asymptomatic children are (ECDC 2020). In addition, because children are more often asymptomatic, they are less likely to be tested, thus causing a potential under-reporting of the prevalence among children. However, a study comprising systematic testing of contacts of confirmed COVID-19 cases in 13 outbreaks in schools in the Norwegian capital region during August-November 2020, found minimal transmission from children (below age 14) to peers and adults (Brandal et al. 2021). In addition, we also present some evidence on effects of school re-openings for fourth graders (age 10) compared to fifth graders (age 11). Because schools re-opened two weeks earlier for the former group, we also compare changes in COVID-19 infection and testing rates among these two groups (and their parents). In accordance with our main results, we find no evidence of increased infection of school re-openings for these two groups either.

The paper proceeds as follows. Section 2 presents relevant background on COVID-19 infection rates and testing in general, and in Norway. Section 3 presents the empirical strategy in this paper, and Section 4 describes the data and presents some summary statistics. In Section 5, we present the results together with a set of robustness checks. Section 6 presents extensions and mechanisms. Section 7 provides a discussion, and Section 8 concludes.

## 2 Background on lockdown and testing in Norway

The first confirmed case of COVID-19 in Norway was registered on February 26, 2020. On March 12, 2020, the government initiated a lockdown to curb the spread of the disease, which included closing all preschools and primary and secondary schools. People were advised to limit the number of close contacts, to meet outside while maintaining physical distance, and to intensify basic hygiene measures. Moreover, all cultural or sporting events and all organised sports were prohibited. Mandated closures of recreational facilities (fitness studios, gyms, swimming pools, etc.), beauty salons (hairdressers, spas, etc.), bars, and restaurants^1^ were enforced. People were advised to avoid all non-essential travel and to limit public transportation, and mandatory quarantine after international travel was enforced. In addition, all health care workers were prohibited from international travel (The Norwegian Directorate of Health 2020).

With very few exceptions, and for the large majority of students, schools remained closed from March 13. Exceptions were largely limited to young children of workers of critical importance for managing the pandemic and basic services, as well as for particularly vulnerable students (2.5 percent of students) or students granted special education (2.5 percent of students).^2^ Unlike primary and secondary schools, universities did not resume in-person teaching in the spring of 2020.^3^

Due to concerns about the consequences of lockdown, particularly for vulnerable children, and to accumulated knowledge of lower COVID-19 infection and morbidity rates among children in general, school closures were among the first measures to be lifted. Preschools and elementary students in grades 1-4 were allowed back in schools starting April 27, while inperson schooling for older students resumed two weeks later (May 11). Schools reopened in a context of low spread of COVID-19 and relatively few other channels for transmission (Telle et al. 2021).

The timing and manner of school re-openings were decided by the national government with very limited geographic variation. In principle, local authorities were allowed some flexibility as to when the reopening was carried out, e.g. government guidelines allowed for regional closures in the case of local outbreaks. That said, the majority of schools reopened in accordance with the legislation: while about 36 percent of elementary schools reported a few days’ delay in order to accommodate the strict infection control guidelines, only a few elementary schools reported further delays. Students quickly returned to school. Among students in elementary school, only 1 percent continued remote learning during the first week (week 20) after reopening.^4^ The reported absence was mostly due to being or having immediate family in the COVID-19 risk group.^5^

When schools were opened after lockdown, extensive public guidelines to curb transmission were given by the health authorities. These measures included self-isolation of sick children and staff, intensification of basic hygiene (hand-washing, frequent cleaning of facilities) and physical distancing. Mask wearing was neither mandated nor encouraged for children. Importantly, a “cohort” system was introduced as the key physical distancing measure. The cohorts consisted of fixed groups of students and employees with limited contact between cohorts while allowing children to socialise within the cohorts (see Johansen et al. (2020) for more details on the guidelines). Cohorts were not recommended for students in high schools because it would excessively limit teaching. Because the current public guidelines were too strict to allow normal teaching, a “traffic light” system was introduced on May 29. Under this system, the intensity of the infection control measures reflected the local infection rates so that the infection control measures were to increase with the infection rates in society.

### Testing

Throughout the period observed in this study, the recommendation was that all persons with symptoms should be tested. While testing was free, test capacity remained limited. Individuals who belonged to prioritized groups took precedence, and on April 20th, before reopening of schools, staff and students were added to the list of prioritized groups.^6^ Frequent testing upon development of symptoms were among the key measures to keep the pandemic under control during re-opening of schools. Teachers were among the groups most frequently tested during the spring 2020 (Magnusson et al. 2021). In short, while a large proportion of cases were always expected to go undetected, the policy of frequent testing means that our data tracks any increase in COVID-19 due to the school re-opening closely.

## 3 Empirical strategy

In this study, we seek to estimate effects of school re-openings on three groups that were directly impacted by school openings, namely students, teachers and parents of school-age children. We implement a difference-in-differences approach comparing infection rates of groups that were directly affected by school openings (students, teachers and parents of high school students, referred to as “in school”) with outcomes of groups that were not directly affected (young adults above school-leaving age, non-teacher professionals and parents of young adults) before and after the re-opening of Norwegian schools.

Schools re-opened in two steps; in-person instructions for the youngest students (pre-school through grade 4) resumed on April 27th, while older students (grades 5-13) were allowed back in school on May 11th. In principle, it is possible to leverage the staggered implementation to identify effects of re-opening on testing and incidence rates. However, the fact that these two dates are so close together leaves us with only a 14 day period to evaluate differential changes in outcomes. Our primary analysis thus focuses on the second step in the re-opening of schools, i.e. May 11th, after which all teachers and students were allowed to resume in-person instruction.^7^ We do not observe the degree to which individual students and teachers were physically present at schools following re-opening. As a consequence, our estimates should not be interpreted as the effects of physical attendance. Rather, we estimate an intention-to-treat (ITT) effect of re-opening schools.

Let *y*_*gt*_ denote the average outcomes of individuals belonging to group *g* in week *t*. Our basic event study regression specification can be written:

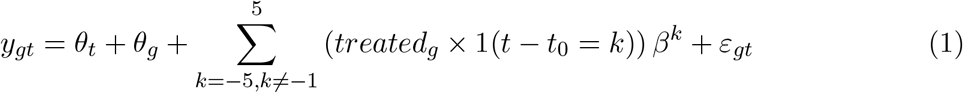

where *θ*_*t*_ and *θ*_*g*_ denote a set of week and group fixed effects.^8^ Our primary parameters of interest are the *β*^*k*^ coefficients, which denote time relative to the week before schools reopened.

We estimate equation (1) on three separate samples. To estimate effects on students, the sample includes teenagers and young adults aged 17-22, where teenagers aged 17-19 (upper secondary school age) are compared to those aged 20-22 (recently graduated). To estimate effects on parents, we compare parents whose youngest child is age 17-19 to those whose youngest child is aged 20-22. Thus, any effects of school re-opening from younger siblings will not contaminate our estimates. In the final sample, teachers employed at primary or secondary schools are compared to other workers in “professional” occupations according to the ISCO-88 classification. See further details on the sample definitions in Section 4.

Our empirical strategy allows for teachers, students and parents to have different COVID-19 incidence levels from their comparison groups. To illustrate, 17-19 year-olds are more likely to still live at home relative to young adults aged 20-22. This difference in living arrangements may affect infection rates independent of whether or not schools are open for in-person instruction. For instance, parents with co-resident teenage children may have higher rates of within-household contagion (and in turn, higher rates of confirmed incidence), relative to parents whose children have all moved out. More generally, the treatment and comparison groups may not have identical outcomes in the absence of treatment. Our econometric model allows for unrestricted level differences between groups (via the inclusion of group fixed effects *θ*_*g*_).

Our key identifying assumption is that each affected group would have trended in parallel with their comparison group, in the absence of schools re-opening. While the parallel trends assumption is inherently untestable, the *β*^*k*^ coefficients for *k <* 0, provides a test for parallel pre-trends. The coefficient path should be close to zero if parallel pre-trends hold. Under this assumption, *β*^*k*^ for *k >*= 0 will capture the causal effect of school opening on COVID-19 outcomes for teachers, students and parents.

We also estimate a set of difference-in-difference models. Our basic difference-in-differences regression specification can be written:

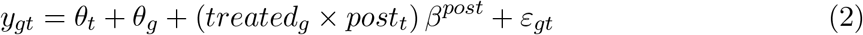

where *θ*_*t*_ and *θ*_*g*_ denote a set of week and group fixed effects (as defined in Equation (1)). Our primary parameter of interest *β*^*post*^ is attached to the interaction term between two indicator variables: *treated*_*g*_, equal to 1 if *g* is directly affected by the school re-opening, and *post*_*t*_, equal to 1 after schools reopen.

To assess robustness with respect to differing trends, we estimate two augmented specifications of Equation (2) that include group-specific time trends. In the first of these specifications, we include a linear time trend interacted with treatment status.

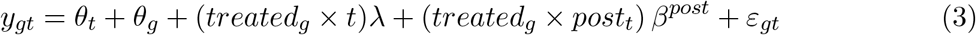

This specification is more flexible as it allows for differential linear trends in outcomes in the treated and comparison groups. At the same time, the estimated paramter *β*^*post*^ could be biased to zero if the group specific time trend estimated in equation (3) effectively absorb some of the effect of reopening. To account for this, we implement a specification where the treatment trends are estimated on pre-reopening data only:

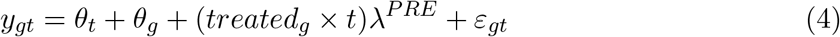

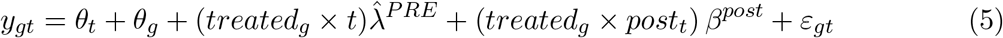

## 4 Data

### 4.1 Beredt C19. The emergency preparedness register for COVID-19

The data applied in this paper are from the Emergency preparedness register for COVID-19 (Beredt C19). The register was established to give the Norwegian Institute of Public Health an ongoing overview and knowledge of the prevalence and consequences of the COVID-19 pandemic in Norway.^9^ It comprises individual-level data from a set of linkable administrative registers, including daily updated records on polymerase chain reaction (PCR) tests and positive COVID-19 cases from the Norwegian Surveillance System for Communicable Diseases (MSIS),^10^ as well as data from the State Register of Employers and Employees (AA register) and the National Population Register. Individuals are linked across the registers and to family members using unique (de-identified) personal identifiers. Consequently, detailed information on demographics (age, gender, county of residence and family members) and employment (industry, occupation, and county of employment) for each individual are merged with data from COVID-19 PCR tests.

Our main outcome variables are (i) the incidence of COVID-19, defined as incidence per 100,000 capita per week, and (ii) test rates, defined as tests per 100,000 capita per week. The number of confirmed cases is an imperfect proxy for the true incidence of COVID-19, because it is likely to reflect variation in the availability of testing as well as variation in underlying incidence.^11^ This is potentially a problem for our analysis if the re-opening of schools shifts the likelihood of being tested differentially among the “in school” group. To assess this possibility, we also analyse the total number of tests.

### 4.2 Sample

We construct three separate regression samples for this study. First, *students* includes all residents aged 17-22, excluding those working as teachers or employed in primary or secondary schools. This sample consists of students in the final years of high school (aged 17-19) and a comparison group consisting of those recently graduated (aged 20-22). Second, *parents* includes the parents of the former sample (*students*). In this sample, outcomes for those where the youngest child is still of school age (17-19) will be compared to those where the youngest child has recently graduated (age 20-22). Finally, *professionals*, includes teachers working at schools defined according to occupation and industry (ISCO-08 codes 23 and NACE codes 85.1-85.3, respectively) and a comparison group of other “professional” workers. Compared to a cross-section of the working-age population, teachers tend to have higher educational attainment. To account for this and other differences in socioeconomic status, we restrict the comparison group to workers in other ISCO-08 category 2 occupations, excluding medical occupations (ISCO-08 code 22). The resulting comparison group, thus, includes science and engineering professionals (ISCO-08 codes 21), business and administration professionals (ISCO-08 codes 24), information and communications technology professionals (ISCO-08 codes 25), and legal, social, and cultural professionals (ISCO-08 codes 26).^12^ We exclude healthcare workers from all samples because they have a substantially different testing regime and exposure to COVID-19 during the sample period.

To reduce computational demands, we collapse the data to the demographic cell-county-week level.^13^ In 2020, there were 11 counties in Norway. Table 1 presents summary statistics of our main estimation samples. Compared to other professionals, teachers are more likely to be female and less likely to live in the capital region (Oslo and Viken counties). Teachers and non-teachers have similar confirmed incidence of COVID-19 during the 12 week window around school re-opening. The two groups differ when it comes to testing, and teachers have an average of 83% more weekly tests around the time of school re-opening. There are relatively small differences in COVID-19 incidence between the two groups of parents, but test rates are higher for parents of older children. For students, infection and test rates are substantially higher for those aged 20-22 compared to those aged 17-19.

**Table 1:**
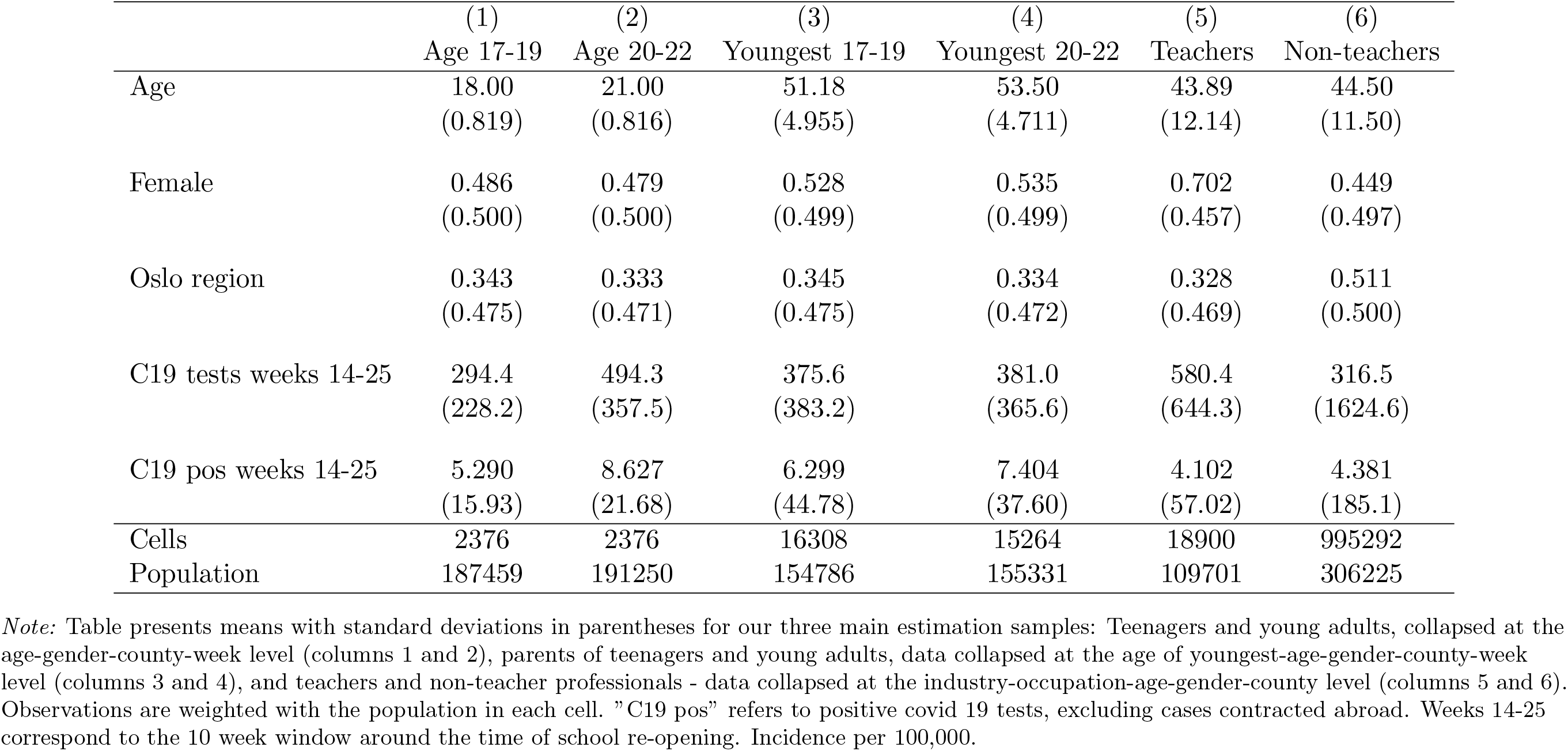
Summary statistics

Trends in incidence for students, parents and teachers are presented in Appendix Figure A1. During most of the period of school closures, teenagers age 17-19 have lower rates of confirmed COVID-19 relative to older young adults. However, incidence in the two groups converges and tracks closely through the reopening week. Incidence among parents and among teachers and other professionals tracks very closely before, during and after the period of school closures. In addition, incidence continues to trend very closely throughout the period for which we have data. Moreover, we find no divergence around the beginning/end of summer vacations, and we see no evidence of divergence during the fall when confirmed incidence rates are higher.

## 5 Results

### 5.1 Event study models

Figure 1 plots the estimated event study models from equation (1) for students, parents, and teachers. For students (Panel a), the figure indicates non-parallel pre-trends. As shown in Figure A1, incidence rates for young adults aged 20-22 and high school age teenagers aged 17-19 tended to converge in the last weeks of school closures. Consistent with these trends, the event study figure finds differential pre-trends that converge several weeks before reopening. Differential pre-trends are less pronounced for parents, and for teachers the pre-trends are close to zero and not statistically significant. The figure gives no indication of significant treatment effects. The estimated event study coefficients do not shift significantly after reopening for either high school students, parents, or teachers (pooled across all grade levels).

**Figure 1:**
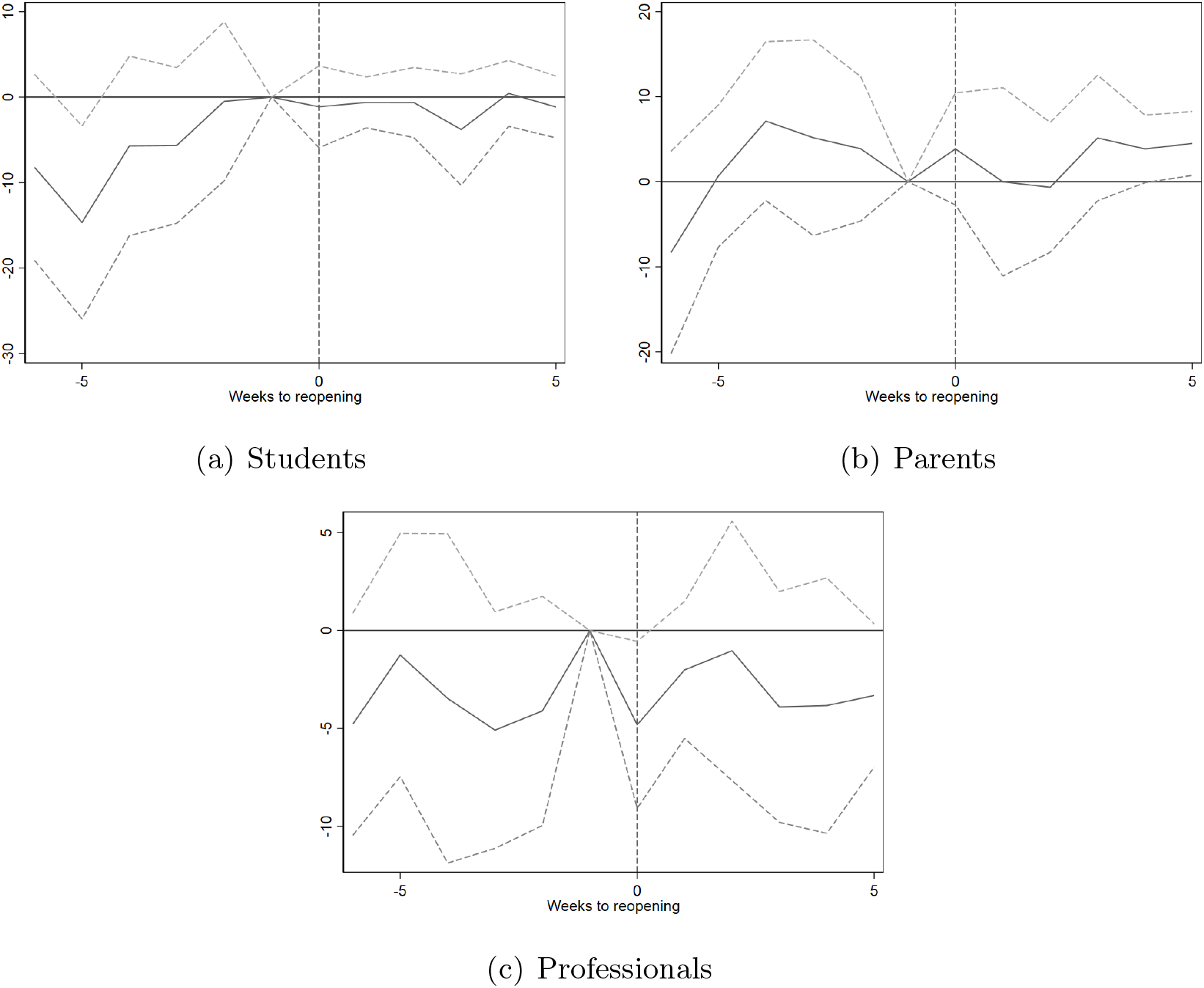
Event study of school reopening on COVID-19 infections by estimation sample *Note:* Figure shows estimates of Equation (1) with 95% confidence intervals. The dependent variable is the number of confirmed positive COVID-19 tests per 100,000 population. Week 19 and the comparison groups as reference. All models include week fixed effects. In Panel (a), models additionally include age fixed effects. In Panel (b), models also include controls for age of youngest child. For Panel (c), models also include controls for industry and occupation. Standard errors are clustered at the county level.

Due to the age gradient in COVID-19 infections, high school students likely pose a larger infection risk than primary and secondary school students. Furthermore, they shift between specialisation groups throughout the week, giving more contact points and potentially facilitating transmissions. To test empirically if contagion is higher in high schools, we estimate effects from teachers in high school and teachers in primary/lower secondary school separately.^14^

Figure 2 shows these separate event study estimates. Among teachers in primary and lower secondary school (left panel), the pattern is much as in the main sample: no significant pre-trends and no infection effects from the re-opening. Among high school teachers, we see a relative increase in infection rates concentrated 2-3 weeks after reopening, as one would expect from the incubation period of COVID-19. However, the effect is estimated with low precision, and the event study coefficients are not statistically significantly different from zero in the post-period. The size of the effect peaks at about 10 infected teachers per 100,000. Reassuringly, the pre-trend does not deviate substantially or significantly from the pre-trend in the comparison group.

**Figure 2:**
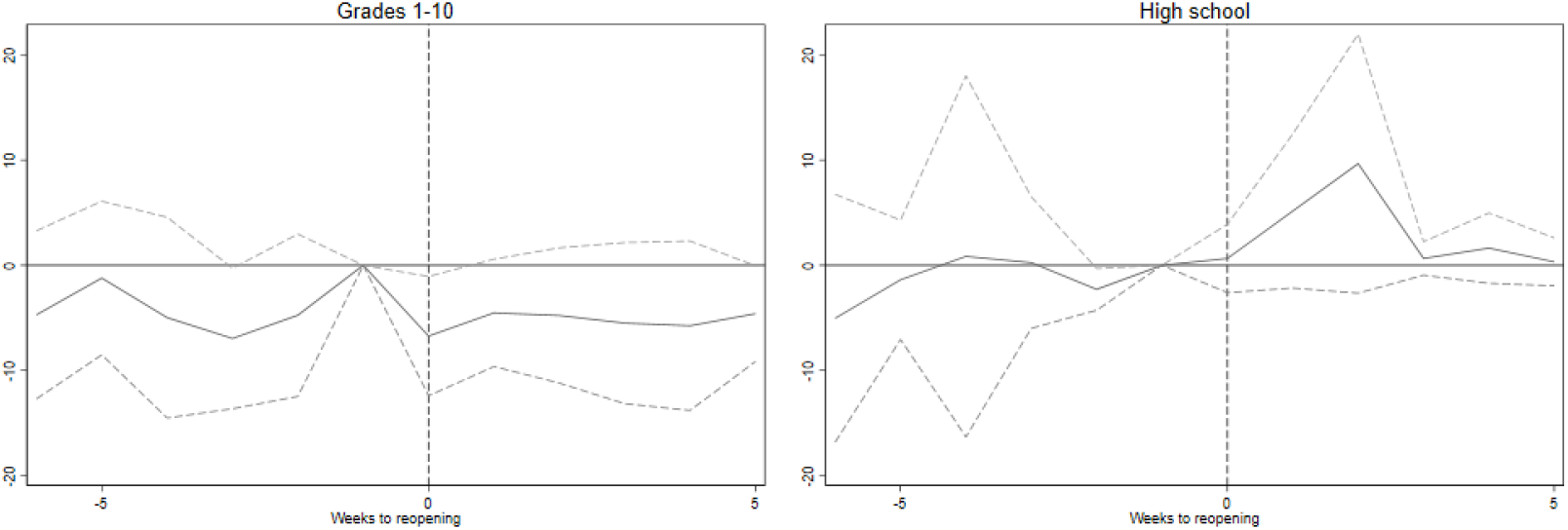
Event study of school reopening on COVID-19 infections by grade level taught *Note:* Figure shows estimates of equation (1) with 95% confidence intervals. The dependent variable is the number of confirmed positive COVID-19 tests per 100,000 population. Week 19 and other ISCO-08 category 2 occupations as reference. Models include week, industry and occupation fixed effects. Standard errors are clustered at the county level.

### 5.2 Difference-in-differences estimates

Table 2 presents estimates from the difference-in-differences specification of equation (2). For teachers, we see no significant effects when grade levels are analysed jointly. The point estimate is close to zero, and the associated 95 percent confidence interval allows us to rule out increases in contagion larger than 2.1 weekly cases per 100,000 teachers. Consistent with the results in our event study models, we find that the effect of re-openings on teachers varies with grade level taught. For teachers at elementary and lower secondary schools (grades 1-10), we estimate a negative, but not statistically significant, effect of reopening. For high school teachers, school re-opening increased COVID-19 infection rates by 4.3 per 100,000 on average in the first 6 weeks following the re-opening. The effect is significant at the 5 percent level.

**Table 2:** Difference-in-difference estimates

|  | (1)<br>Students | (2)<br>Parents | (3)<br>Teachers | (4)<br>Teachers |
| --- | --- | --- | --- | --- |
| *Panel A: Baseline* |  |  |  |  |
| Reopening x treated | 4.64***<br>(1.67) | 1.36<br>(1.86) | -0.0358<br>(1.09) |  |
| Reopening x grade 1-10 |  |  |  | -1.54<br>(1.34) |
| Reopening x high school |  |  |  | 4.28**<br>(1.76) |
| *Panel B: Group lin trends* |  |  |  |  |
| Reopening x treated | -2.52<br>(4.21) | -4.63<br>(3.12) | -1.14<br>(1.32) |  |
| Reopening x grade 1-10 |  |  |  | -2.65**<br>(1.33) |
| Reopening x high school |  |  |  | 3.17<br>(2.32) |
| *Panel C: Group linear pre-trends* |  |  |  |  |
| Reopening x treated | -8.11***<br>(1.67) | -1.69<br>(1.86) | -0.514<br>(1.09) |  |
| Reopening x grade 1-10 |  |  |  | -2.02<br>(1.34) |
| Reopening x high school |  |  |  | 3.80**<br>(1.76) |

*Note:* Difference-in-differences estimates from the specification in equation (2). All models include week, industry, and occupation fixed effects. In Panel (B), models additionally include group linear trends. In Panel (C), models additionally include group linear trends with the coefficient estimated on pre-period data only (equation (5)). Standard errors are clustered at the county level. \* $p < 0.10$ , \*\* $p < 0.05$ , \*\*\* $p < 0.01$

We find no statistically significant effects for parents. For students, there is a significant increase in confirmed cases following reopening. Given the differential pre-trends presented in Figure 1, this estimate should, however, not be given a causal interpretation.

### 5.3 Robustness

#### 5.3.1 Differential trends

To assess the robustness of the difference-in-differences estimates with respect to differential pre-trends, we estimate an augmented specification with a linear time trend interacted with treatment status. Results from this exercise are shown in Panels B and C of Table 2. The corresponding event study plots can be found in Appendix Figure A5.

Panel B presents estimates from the in Equation (3), where treatment status is interacted with a linear time trend; this specification substantially reduces the precision of the estimates. The point estimates for students and parents switch signs, and the effect on students is no longer statistically significant. With group-specific linear time trends, we now estimate a significant negative effect of re-opening on teachers in grades 1-10. Taken at face value, this would suggest that re-opening schools reduced incidence in this group. However, the event study plots in Figure 2 suggest that this negative effect likely reflects diverging trends, as there was no corresponding drop in incidence. Adding group linear time trends, the estimated effect on high school teachers is no longer significantly different from zero.

Meanwhile, the specification underlying the estimates in Panel B is known to bias estimates in the presence of time-varying treatment effects, see, e.g. Borusyak & Jaravel (2017). To account for this, panel C presents estimates from Equation (5), where the coefficient on group-specific time trends is obtained from an auxiliary model using only pre-intervention data. In this specification, we now obtain a significant negative effect of re-opening on incidence for secondary school students. Moreover, in this specification, the effect on high school teachers is once again statistically significant, with a point estimate similar to the effect obtained from our baseline model.

To summarize, the estimated effect of reopening on contagion rates for students appears to be sensitive to differential trends; consistent with the differential pretrends evident in the event study plots of Figure 1. For parents and teachers overall, we find no significant impact of re-opening in either specification. In contrast, the effect for high school teachers appears more robust to the inclusion of group-specific trends.

#### 5.3.2 Treatment groups

Our models find no significant effects of school re-openings on teachers pooled across elementary and secondary school levels. However, there could still be effects on people working in non-teacher occupations, e.g. support staff working in schools. Similarly, there may be effects on kindergarten workers. To assess this, we estimate additional event study models estimating treatment effects on two additional groups: (1) all workers employed in schools and (2) all workers employed in schools or kindergartens, in teacher or non-teacher occupations. In these models, the comparison group consists of all workers. Results from these models, presented in Appendix Figure A2, find no indication that the re-opening of schools and kindergartens increased the number confirmed COVID-19 cases among treated workers more broadly defined.

## 6 Extensions and mechanisms

### 6.1 Effects on primary school students

Our main models estimate effects of re-opening on upper secondary school students and their parents, using young adults just above school-leaving age as a control group. The re-opening of schools was implemented step-wise, with the youngest students (preschool through grade 4) returning to school 2 weeks before older students (grades 5-13). To assess whether the reopening of schools for the youngest students affected COVID-19 infections, we have estimated a set of event study models of confirmed COVID-19 incidence among fourth grade students and their parents, using 5th graders and their parents as a natural comparison group.^15^

Results from this exercise are presented in Figure 3. Due to the brief interval between the reopening of schools for the two age groups, we only have a 2-week period to assess the effects of reopening. With that caveat, we note that the results in Figure 3 are consistent with the conclusions from our main specifications. We find no indication that the earlier re-opening of schools for the younger children increased contagion rates among primary school students or their parents.

**Figure 3:**
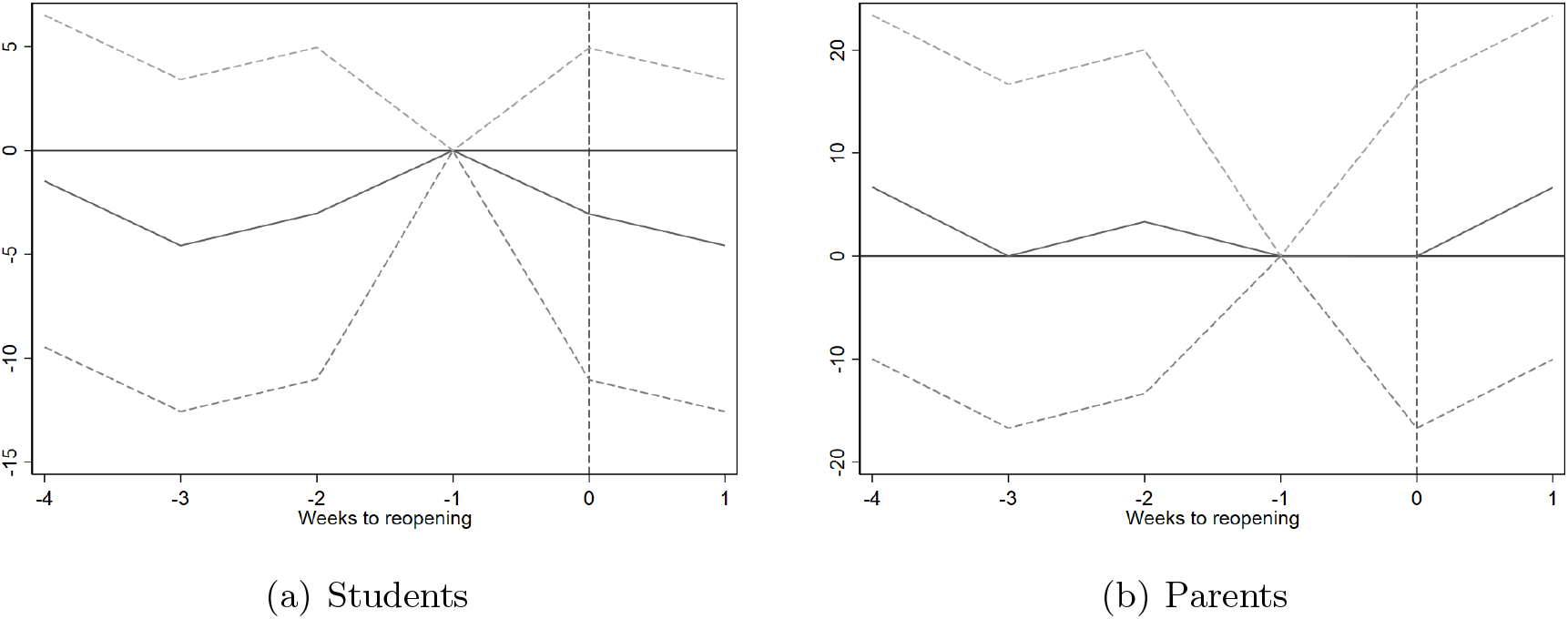
Event study of school reopening on COVID-19 infections by fourth versus fifth grade students and their parents *Note:* Figure shows estimates of equation (1) with 95% confidence intervals. The dependent variable is the number of confirmed positive COVID-19 tests per 100,000 population. Week 17 and fifth graders as reference. All models include week fixed effects. In Panel (a), models additionally include age fixed effects. In Panel (b), models include controls for age of youngest child. Standard errors are clustered at the county level.

### 6.2 Strength of effects and the infection level in society

One reasons for our modest effects, could be that infection levels among students are very low, or simply zero. Due to the exponential nature of infection, effects may be substantially stronger in contexts with even modestly higher infection. For relevance to contexts with higher overall infection, we explore effects in two contexts of relatively higher infection within our sample.

During the first wave of the pandemic, the confirmed incidence of COVID-19 was significantly higher in the Oslo region (Oslo and Viken counties) relative to the rest of the country. In the 12-week window around re-opening, average weekly incidence was more than five times higher among high school age teenagers residing in the Oslo region relative to teenagers in the same age group in other counties (10.6 vs 2.2 per 100,000).

To address this question, we estimated a triple difference model building on the specification in Equation (2) which includes a triple interaction term capturing the differential impact of school re-openings on treated groups in the Oslo region.

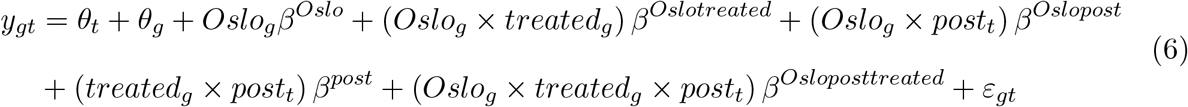

This triple difference specification puts great demands on the data, and given the low baseline incidence, we are likely underpowered in estimating all relevant interaction terms. With that caveat, results from this model are presented in Table 3.

**Table 3:** Triple difference estimates Oslo region

|  | (1)<br>Students | (2)<br>Parents | (3)<br>Teachers | (4)<br>Teachers |
| --- | --- | --- | --- | --- |
| Reopening x treated | 4.56*<br>(2.53) | 2.15<br>(2.03) | -0.0592<br>(0.689) |  |
| Reopening x treated x Oslo | 0.552<br>(2.57) | -1.94<br>(4.22) | -2.40**<br>(1.08) |  |
| Reopening x grade 1-10 |  |  |  | -0.785<br>(1.07) |
| Reopening x high school |  |  |  | 2.03<br>(1.32) |
| Reopening x grade 1-10 x Oslo |  |  |  | -4.80***<br>(1.08) |
| Reopening x high school x Oslo |  |  |  | 4.39<br>(2.88) |

*Note:* Difference-in-differences-in-differences estimates from equation (6). Standard errors are clustered at the county level. \* $p < 0.10$ , \*\* $p < 0.05$ , \*\*\* $p < 0.01$

The estimated models do not indicate any additional increases in incidence among treated groups in the Oslo region following reopening. The models find significant negative interaction terms for teachers in the Oslo region following re-opening, driven by teachers in grades 1-10. However, due to the numerous statistical tests performed and absence of consistent evidence for a negative effect in this group, we are hesitant to put too much weight on these estimates. Moreover, Appendix Figure A3 presents event study models estimated separately for the Oslo region versus other counties. The confidence intervals are wider for the Oslo sample, likely reflecting the smaller sample size. Meanwhile, we find similar point estimates for the Oslo region sample and the other counties. This pattern holds for both students, parents, and teachers. To summarize, we find no indication that COVID-19 rates increase at re-opening in the relatively high incidence Oslo region.

### 6.3 Testing

The weekly rate of confirmed COVID-19 is an imperfect proxy for the true underlying incidence of the virus, as we only capture cases among patients who are tested. Put differently, there is an unknown number of undiagnosed infections that are missing from our data. This missing data issue has two consequences for our analyses. First, incomplete testing may have implications for interpretation of our effect sizes. While our baseline estimates allow us to rule out post-reopening increases in confirmed cases among teachers greater than 4 weekly cases per 100,000 teachers, we cannot rule out additional increases in undiagnosed cases. We note that our measure is somewhat more robust for symptomatic cases, because persons with symptoms of COVID-19 were encouraged to get tested. Second, school closures and re-openings may have effects on the probability that affected groups get tested for the virus. If students or teachers are more likely to get tested when schools are open, this could show up in our models as an increase in confirmed cases even if there is no effect of reopening on underlying prevalence.

Plotting the trends in testing indicate that testing patterns differ between treatment and comparison groups (see Appendix Figure A4). In particular, the rate of testing rose sharply for young adults aged 20-22 relative to 17-19 year-olds at the time of school closures and remained higher throughout the lock-down and re-opening (Panel A). Starting mid-lock-down, test rates among teachers rose sharply compared to other professionals (Panel C). Parents have similar trends in testing throughout lock-down and the weeks that followed (Panel B). Estimated event study models of testing fail to show a discontinuous shift at the time of school reopening for parents (see Appendix Figure A6, Panel B). In other words, the reopening of schools did not increase testing among parents, the only group for which we have credible causal evidence.

## 7 Discussion

The decision of whether or not to resume in-person schooling has been a key question for policymakers in the handling of the COVID-19 pandemic. In this paper, we use comprehensive register data to analyze what happened to incidence when schools were reopened in Norway following the initial national lockdown during the Spring of 2020. Using a difference-indifference approach, we find no evidence that school re-openings significantly increased the incidence of COVID-19 among affected students, parents, or teachers pooled across grade levels. These results show that at low levels of contagion, about 5 weekly cases per 100,000, reopening of schools can be done safely, provided that open schools are combined with other social distancing measures and relevant safety protocols are implemented in the schools.

While these results are encouraging for stakeholders wishing to reopen schools, these findings come with several caveats. For one, while our analysis focuses on highly exposed groups such as teachers, students and parents, school reopenings could have indirect effects on the comparison groups as well, e.g. if re-opening increases contagion rates in the wider community. While this kind of community spillover seems unlikely given our lack of significant effects on highly exposed groups, they cannot be ruled out. In the presence of such spillovers, our estimates reflect the differential effect of re-opening on highly exposed groups, rather than the overall effects of school re-opening on COVID-19 incidence.

As a result of the safety protocols implemented by schools following re-opening, students and teachers may have spent less time physically present at school compared to before the pandemic. While more than 99% of elementary school students resumed in-person instruction immediately after re-opening, we do not have access to the corresponding numbers for secondary school students or teachers. Reduced physical presence following re-opening does not affect the validity of our empirical design, however it is relevant for the interpretation of our estimates: If we consider the treatment of interest to be the re-opening of schools with full physical presence, our estimated effects should be interpreted as intention-to-treat.

The low-incidence context of this paper implies that our statistical power will be limited. While our point estimates indicate small increases in incidence, the low baseline rates and the resulting lack of statistical power means we are unable to rule out large relative increases. The upper bound of the 95% confidence intervals associated with the estimates in Table 2 imply that incidence could increase with as much as 33% for teachers, 49% for parents, and 90% for students relative to their respective baseline incidence rates in the 6 weeks prior to reopening.

More generally, the low incidence setting prompts questions about how generalizable these findings may be. Schools were re-opened at a time of low rates of incidence and hospitalization. Average weekly incidence in the 5-week period immediately after re-opening was only 1.83 per 100,000 population. These rates are also low relative to the rates observed in other European countries at this time (see Appendix B for a more comprehensive analysis and discussion of the international context). While the early summer of 2020 was a time when several European countries saw low rates of COVID-19 infections (see Appendix Figure B1), a majority of these countries saw significantly higher rates than what we observe in our data. However, we note that on a relative scale, our estimates are comparable to those found by Vlachos et al. (2021) for Sweden for parents and high school teachers, albeit not for teachers of younger children.^16^

When making decisions on in-person schooling, policymakers must weigh the benefits of in-person instruction against the costs in terms of higher incidence of COVID-19. Our findings suggest that when safety protocols are put in place in schools and communities, schools can be safely reopened when transmission rates are sufficiently low. A pressing question then is how we should define the threshold for “sufficiently low”. We see no disproportionally stronger effects in the Oslo region, where contagion is higher. It should be noted that infection rates are relatively low also in this region, at about 20 cases per 100,000. Given the caveats outlined above, we are hesitant to extrapolate beyond this range.

## 8 Conclusion

The findings from this paper add to an extremely limited literature on the safety of re-opening schools – and, consequently, keeping schools open – in a context of relatively low COVID-19 contagion. Using detailed register data covering the universe of Norwegian residents, we compare infection rates for groups with different exposure to in-person schooling across the re-opening of schools during the 2020 spring lock-down in Norway. We show that infection rates among teachers in general are similar to those of comparable professions both before and after the re-opening of schools. The same holds for adolescents attending high school compared to their recently graduated counterparts and for the parents of these two groups of teenagers and young adults. We do find a slight increase in infection rates for high school teachers compared to other professionals. However, this increase was not statistically significant across specifications, and the number of extra cases was small (about 4 per 100,000).

So far, the limited evidence in this field has found that in-person schooling contributes little to the spread of COVID-19. The results in this paper are in line with those findings. The majority of studies are based on infection numbers from spring or summer 2020. As the fall and winter of 2020-2021 has seen a surge in infections across Europe and the US, with a larger share of infections among adolescents and young adults, an important next step is to assess infections from in-person schooling under these higher infection rates.

## Data Availability

Sensitive data kept in repository

## Appendix A

**Figure A1:**
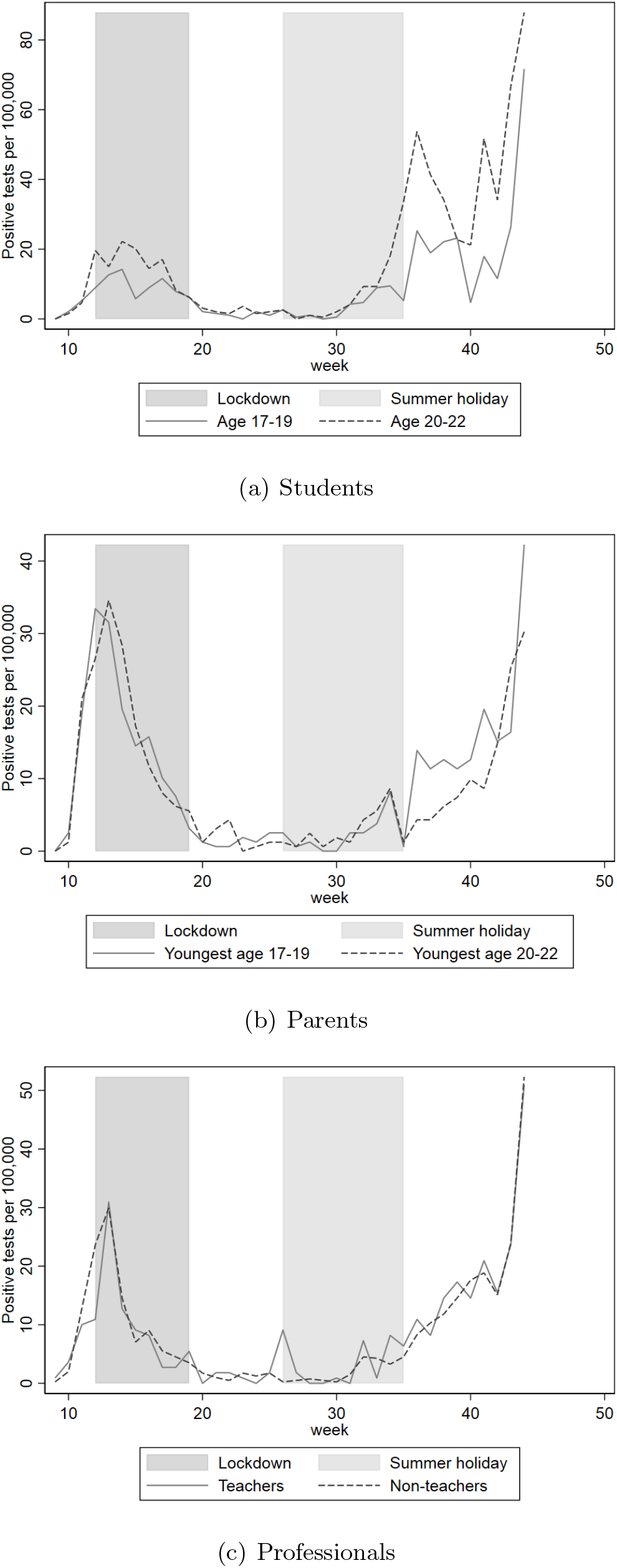
Positive COVID-19 test rates by estimation sample *Note:* Figure shows weekly rates of positive COVID-19 tests for students by age group (high school age versus above high school age), for parents by age groups of youngest child (high school age versus above high school age) and for teachers versus other category 2 ISCO-08 occupations.

**Figure A2:**
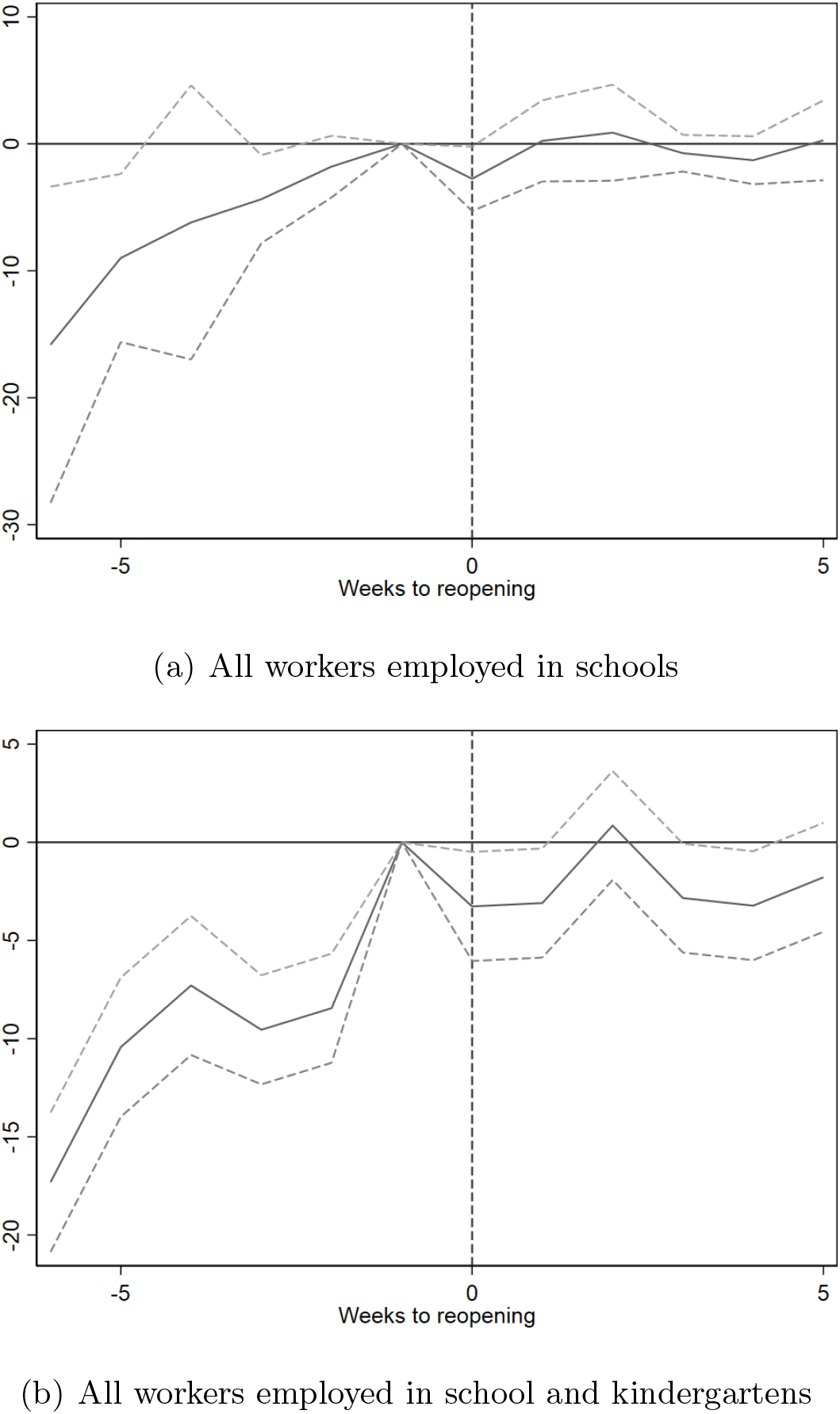
Event study of school reopening on COVID-19 infections on all school staff *Note:* Figure shows estimates of equation (1) with 95% confidence intervals. The dependent variable is the number of confirmed positive COVID-19 tests per 100,000 population. Models include occupation, industry and week fixed effects. The comparison group includes all workers not employed in schools or kindergartens. Week 19 and the comparison groups as reference. Standard errors are clustered at the county level.

**Figure A3:**
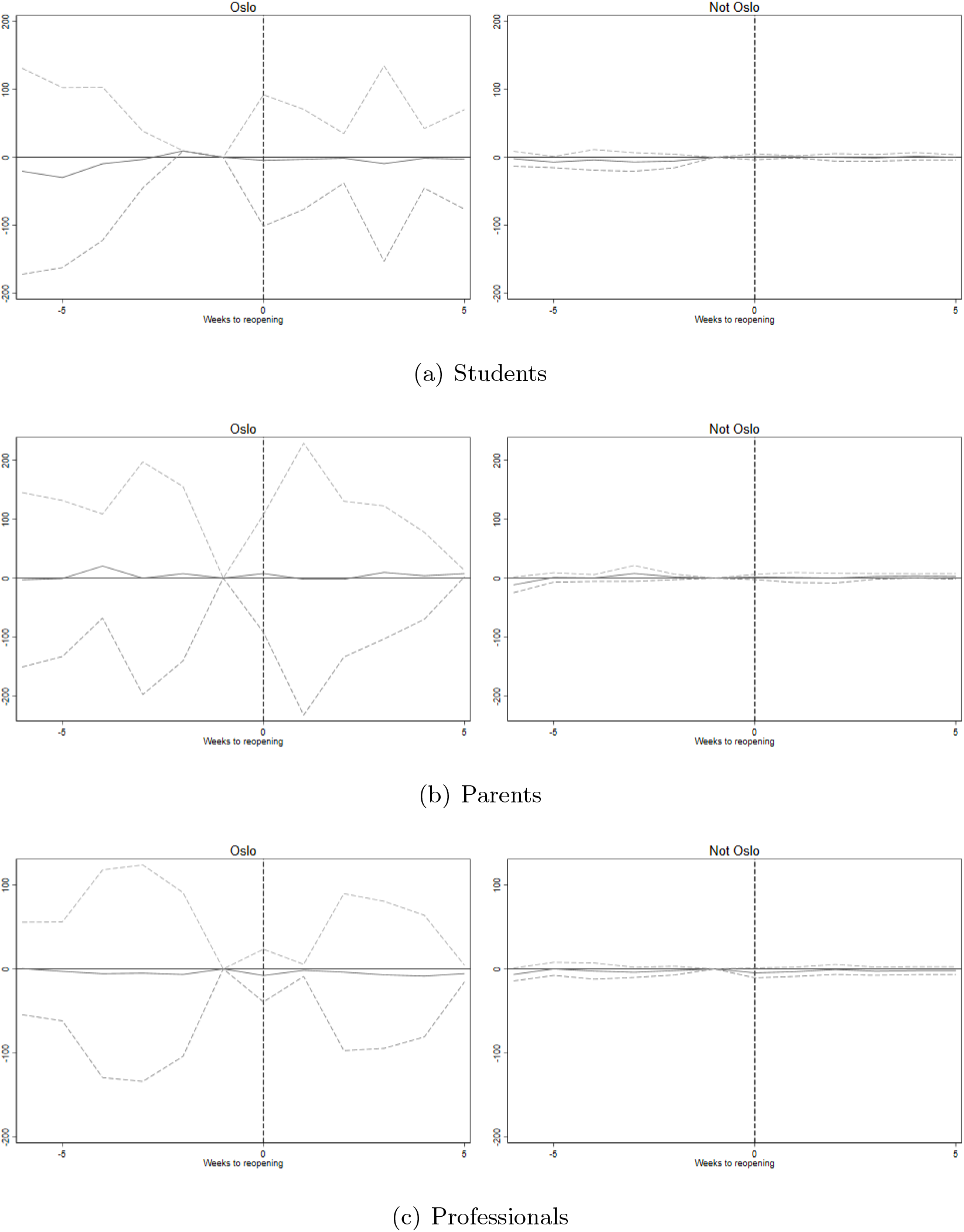
Event study of school reopening on COVID-19 infections in the Oslo area versus other counties *Note:* Figure shows estimates of Equation (1) with 95% confidence intervals. The dependent variable is the number of confirmed positive COVID-19 tests per 100,000 population. Week 19 and the comparison groups as reference. All models include week fixed effects. In Panel (a), models additionally include age fixed effects. In Panel (b), models include controls for age of youngest child. In Panel (c), models include controls for industry and occupation. Standard errors are clustered at the county level.

**Figure A4:**
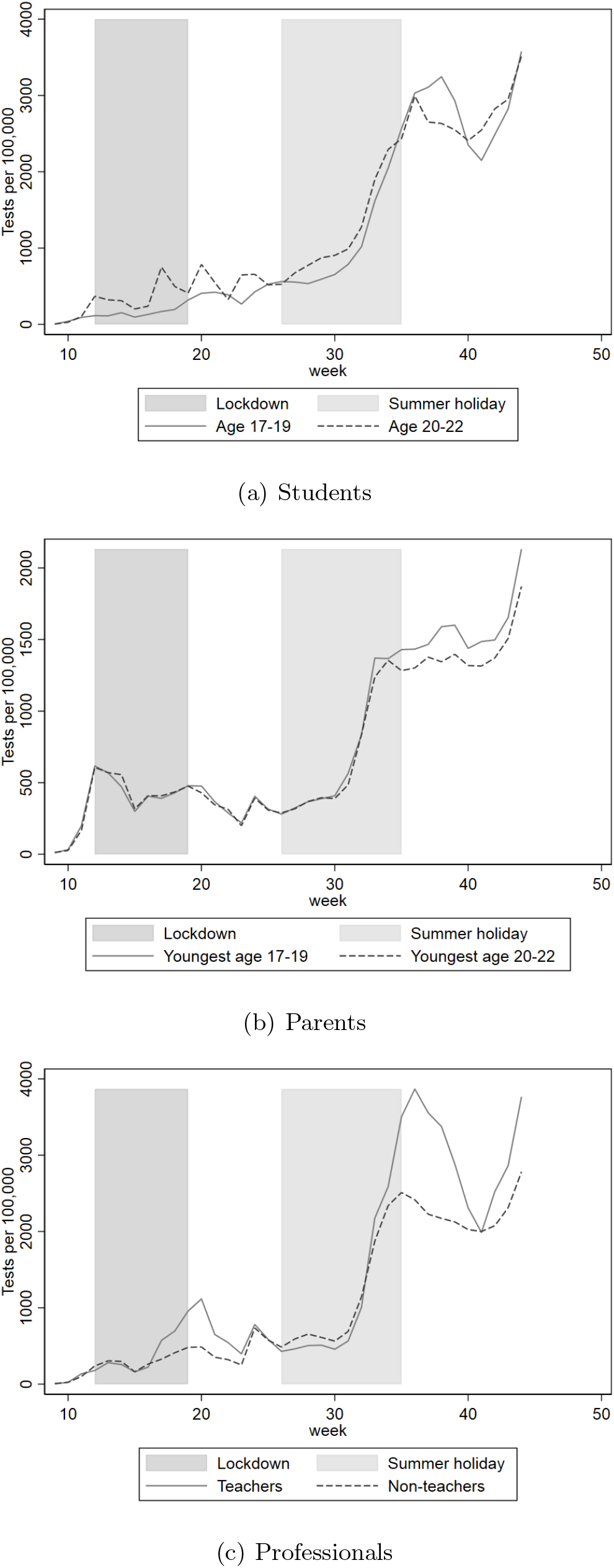
Trends in COVID-19 tests by estimation sample *Note:* Figure shows trends in weekly rate of COVID-19 tests for students by age (high school age versus above high school age), parents (youngest child in high school age versus youngest child above high school age) and teachers versus other professionals (category 2 ISCO-08 occupations).

**Figure A5:**
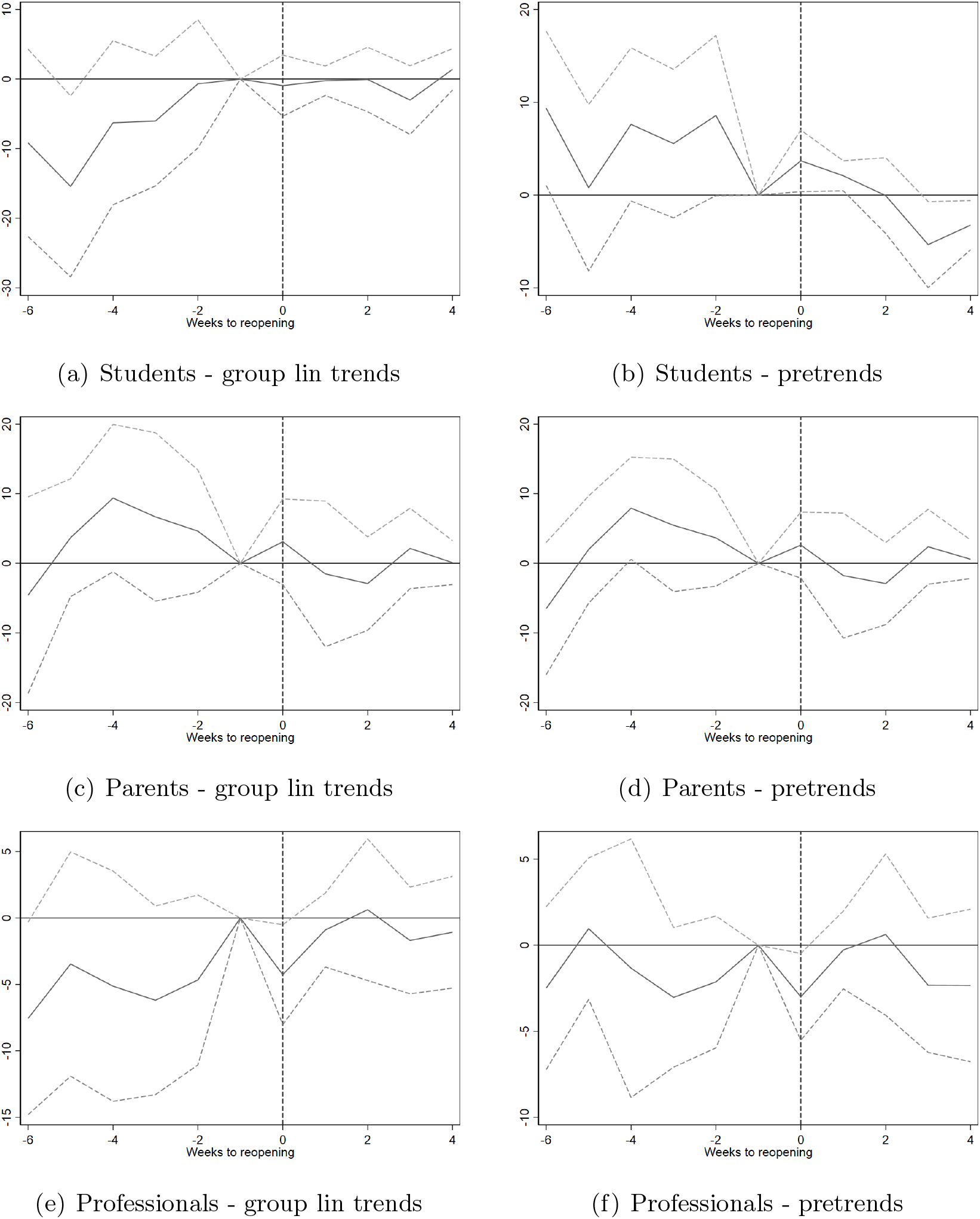
Robustness: event studies with group specific trends *Note:* Figure shows estimates of Equation (1) with 95% confidence intervals. The dependent variable is the number of confirmed positive COVID-19 tests per 100,000 population. Week 19 and the comparison groups as reference. All models include week fixed effects. In Panel (a), models additionally include age fixed effects. In Panel (b), models also include controls for age of youngest child. For Panel (c), models also include controls for industry and occupation. Standard errors are clustered at the county level.

**Figure A6:**
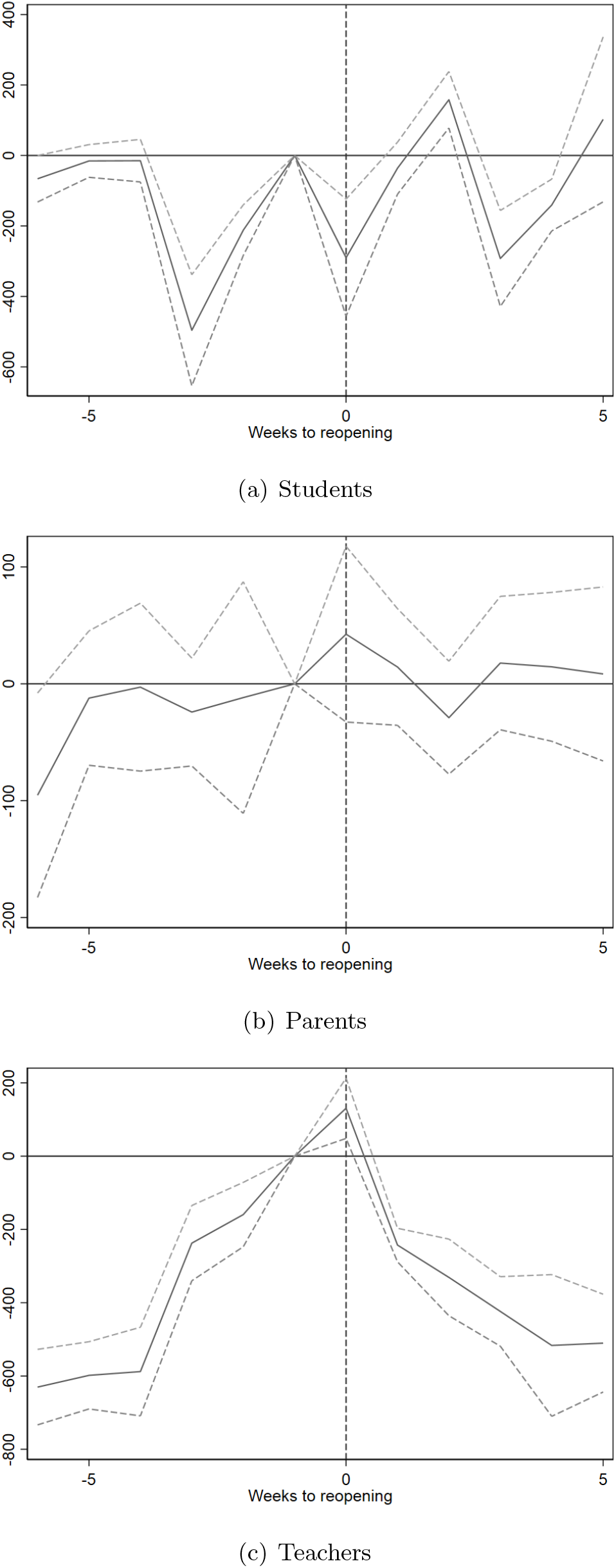
Event study of school reopening on COVID-19 tests *Note:* Figure shows estimates of equation (1) with 95% confidence intervals. The dependent variable is the number of COVID-19 tests per 100,000 population. Week 19 and the comparison groups as reference. All models include week fixed effects. In Panel (a), models additionally include age fixed effects. In Panel (b), models include controls for age of youngest child. In Panel (c), models include controls for industry and occupation. Standard errors are clustered at the county level.

## Appendix B: International context

The timing of the reopening corresponds with the low point of the pandemic in Norway. In particular, the early summer of 2020 was a time when several European countries saw low rates of COVID-19 infections (see Figure B1).

In Norway, the average incidence in the post-reopening weeks (weeks 20-25) was only 1.83 per 100,000 population. Figure B2 shows incidence by country during this period for 45 European countries. Out of these 45 countries, only 9 had lower rates of confirmed incidence than Norway.

In the weeks between March 2nd and December 27th, Norway had weekly incidence rates *rate*_*week*_ *<* 1.83 for 6 out of 43 weeks. Of 44 other European countries, 15 experienced 6 or more weeks of low incidence at or below this level during 2020 (see Figure B3).

**Figure B1:**
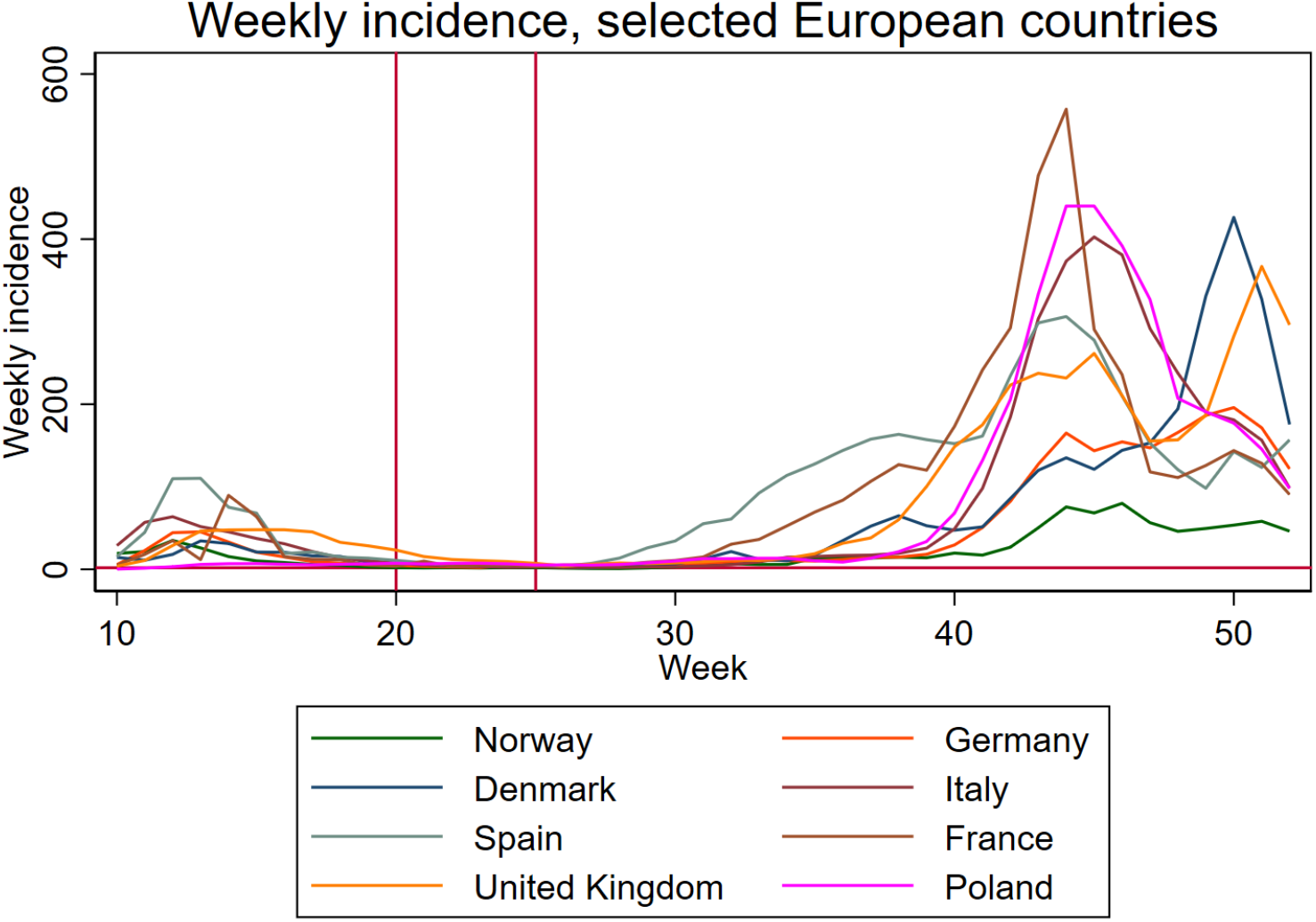
International context *Note:* Figure plots average weekly incidence for selected European countries. Source: Our World In Data.

**Figure B2:**
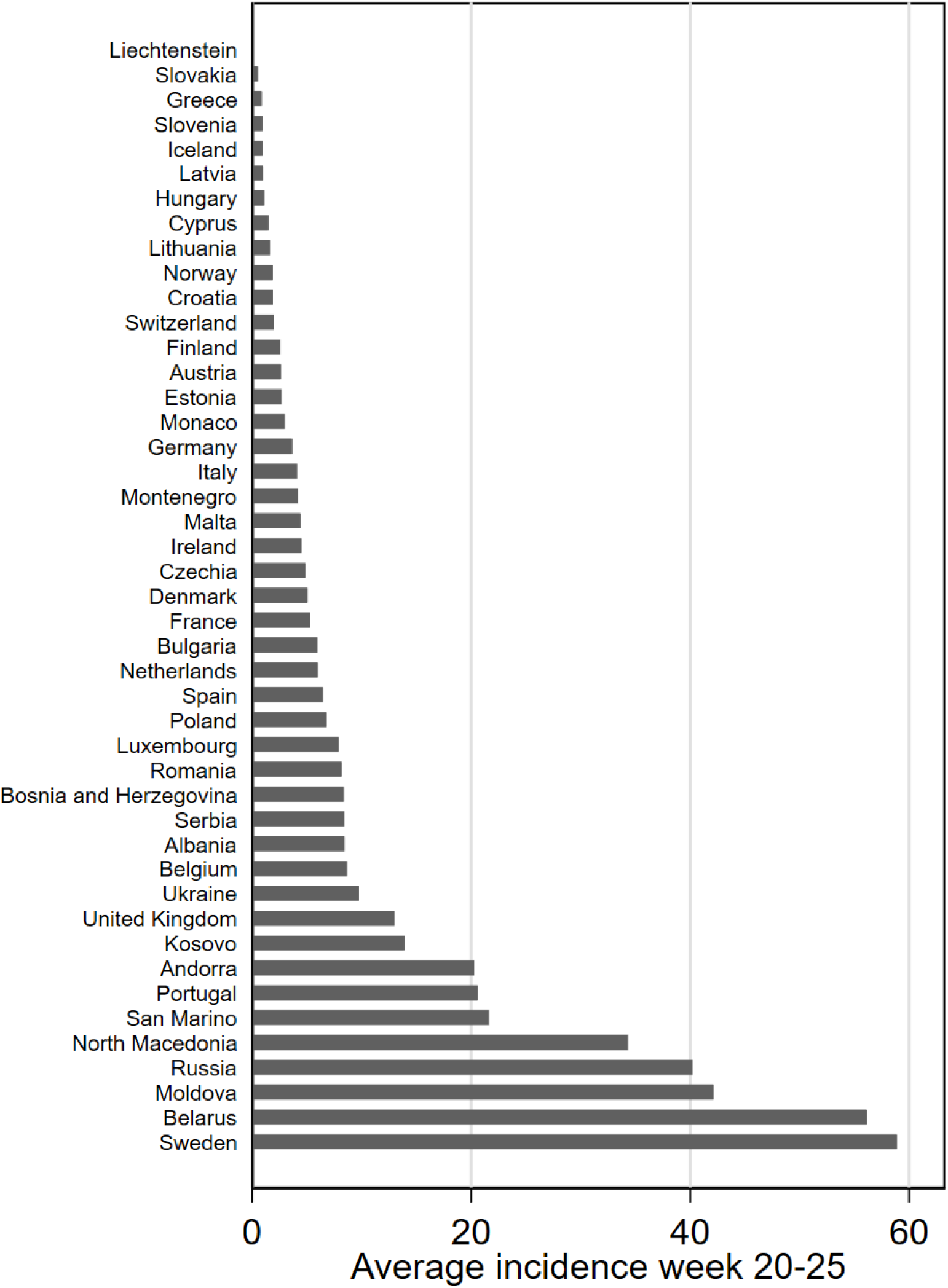
International context *Note:* Figure plots average weekly incidence for selected European countries. Source: Our World In Data.

**Figure B3:**
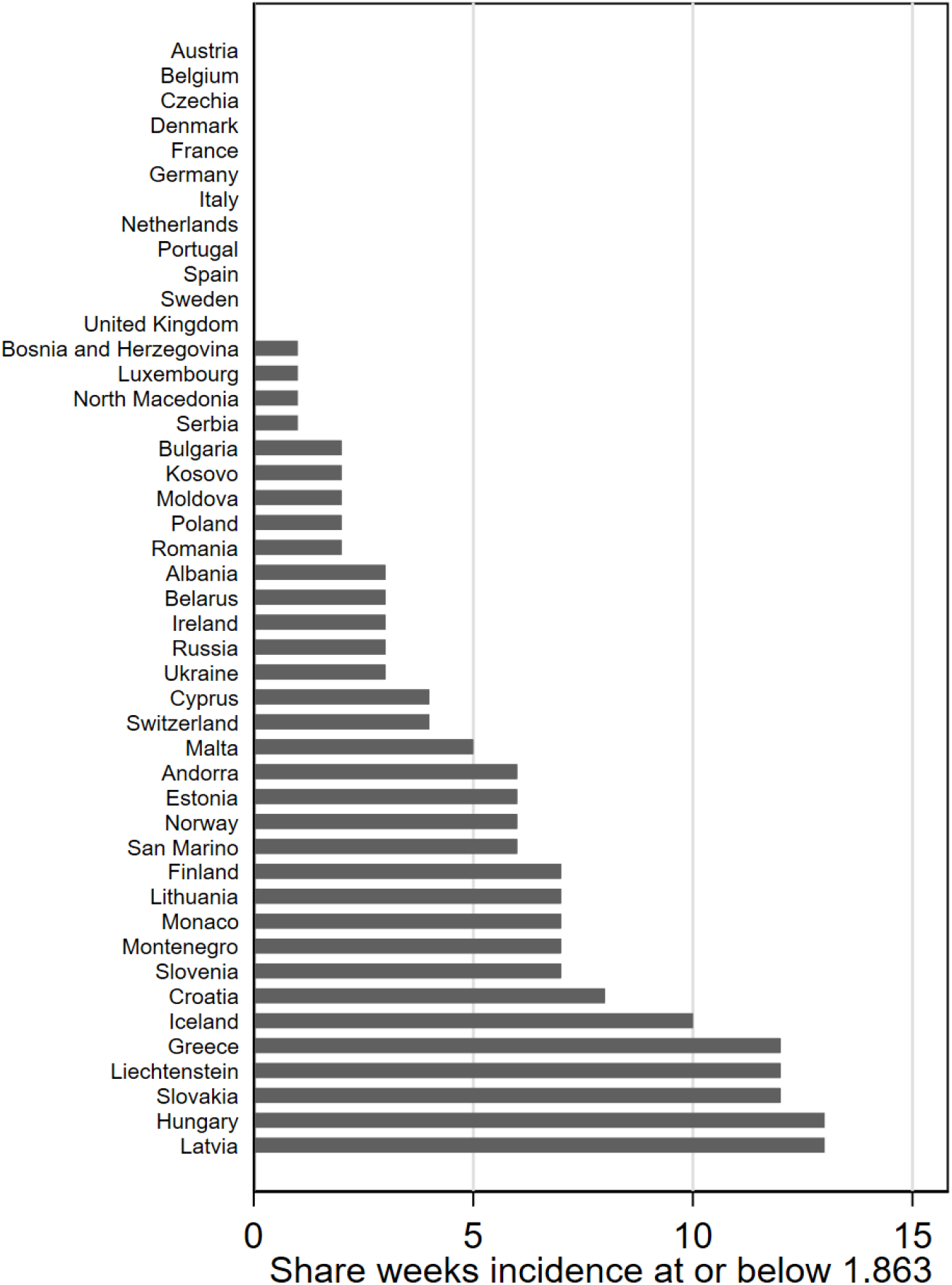
International context *Note:* Figure shows number of weeks in 2020 when weekly incidence was at or below 1.83 per 100,000 residents (the average weekly incidence in the post-reopening period). Source: Our World In Data.

## Footnotes

1 An exception was given to restaurants where physical distance in excess of 1 meter could be maintained.

2 These numbers are from elementary schools (The Norwegian Directorate for Education and Training 2020). Similar numbers from high schools are not available.

3 Exceptions were limited to final year students in performing arts, media and design, science and technology and healthcare who were planning to complete their studies in the spring of 2020, and who required access to campus infrastructure such as labs and practice rooms to complete their degrees. In the 20-22 age group, this is presumably a small number of students.

4 In week 24, this number had fallen to 0.5 percent.

5 The figures presented here are from elementary school re-openings presented in a report by The Norwegian Directorate for Education and Training (2020). There are no such public reports for high schools, however, correspondence with local authorities supports that high schools opened in accordance with legislation and that the majority of students were present, at least to some degree, each week.

6 The description of testing is based on logs of testing guidelines provided to the researchers by the Norwegian Institute of Public Health.

7 For completeness, we have analyzed the impact of the first stage re-opening on 4th grade students and their parents, using 5th graders as a comparison group. Results from these models, presented in Section 6, are consistent with our main findings.

8 For students, the group fixed effects are a set of age dummies; for parents, the group fixed effects are a set of dummies for age of youngest child; for teachers, the model includes occupation and industry fixed effects.

9 The authors obtained access to the data by their roles as analysts in the Beredt C19. The data are kept in a repository administered by the Norwegian Institute of Public Health and is only accessible to other analysts in Beredt C19. The study has not been pre-registered.

10 New cases that test positive by COVID-19 PCR tests are legally subject to notification to MSIS without delay.

11 See Depalo (2021) for a discussion of issues with measuring incidence (and mortality) using administrative data.

12 In one of the robustness specifications, a wider range of workers are applied as the comparison group to a wider definition of school workers.

13 For students, data are collapsed by week, age, county and gender. For parents, data are collapsed by age of youngest child, week, own age (in 5-year bins), county and gender. For teachers, data are collapsed by occupation, industry, week, own age (in 5-year bins), county and gender.

14 The data do not allow us to distinguish between primary and lower secondary schools.

15 As before, parents are grouped by age of youngest child, to avoid spillovers. While we have data on whether teachers are employed in primary or secondary school, we cannot identify precisely which grade levels each teacher is working with. As a consequence, we are unable to leverage this variation to identify effects on teachers.

16 The point estimate of 1.36 new cases per 100,000 corresponds to a 12% increase, comparable to the 20% increase for Swedish parents. For teachers overall, our estimate deviates from that of Vlachos et al. (2021) with an upper bound of 33% increase. For high school teachers, the point estimate of 4.36 cases per 100,000 gives a 61% increase relative to the mean for teachers, again comparable to the Swedish case.

